# Exploring surveillance data biases when estimating the reproduction number: with insights into subpopulation transmission of Covid-19 in England

**DOI:** 10.1101/2020.10.18.20214585

**Authors:** Katharine Sherratt, Sam Abbott, Sophie R Meakin, Joel Hellewell, James D Munday, Nikos Bosse, CMMID Covid-19 working group, Mark Jit, Sebastian Funk

## Abstract

The time-varying reproduction number (*R*_*t*_: the average number secondary infections caused by each infected person) may be used to assess changes in transmission potential during an epidemic. While new infections are not usually observed directly, they can be estimated from data. However, data may be delayed and potentially biased. We investigated the sensitivity of *R*_*t*_ estimates to different data sources representing Covid-19 in England, and we explored how this sensitivity could track epidemic dynamics in population sub-groups.

We sourced public data on test-positive cases, hospital admissions, and deaths with confirmed Covid-19 in seven regions of England over March through August 2020. We estimated *R*_*t*_ using a model that mapped unobserved infections to each data source. We then compared differences in *R*_*t*_ with the demographic and social context of surveillance data over time.

Our estimates of transmission potential varied for each data source, with the relative inconsistency of estimates varying across regions and over time. *R*_*t*_ estimates based on hospital admissions and deaths were more spatio-temporally synchronous than when compared to estimates from all test-positives. We found these differences may be linked to biased representations of subpopulations in each data source. These included spatially clustered testing, and where outbreaks in hospitals, care homes, and young age groups reflected the link between age and severity of disease.

We highlight that policy makers could better target interventions by considering the source populations of *R*_*t*_ estimates. Further work should clarify the best way to combine and interpret *R*_*t*_ estimates from different data sources based on the desired use.

## Background

Within six months of its emergence in late 2019, the novel coronavirus SARS-CoV-2 had caused over six million cases of disease (Covid-19) worldwide (1). Its rapid initial spread and high death rate prompted global policy interventions to prevent continued transmission, with widespread temporary bans on social interaction outside the household (2). Introducing and adjusting such policy measures depends on a judgement in balancing continued transmission potential with the multidimensional consequences of interventions. It is therefore critical to inform the implementation of policy measures with a clear and timely understanding of ongoing epidemic dynamics (3,4).

In principle, transmission could be tracked by directly recording all new infections. In practice, real-time monitoring of the Covid-19 epidemic relies on surveillance of indicators that are subject to different levels of bias and delay. In England, widely available surveillance data across the population includes: 1) the number of positive tests, biased by changing test availability and practice, and delayed by the time from infection to symptom onset (if testing is symptom-based), from symptom onset to a decision to be tested and from test to test result; 2) the number of new hospital admissions, biased by differential severity that triggers care seeking and hospitalisation, and additionally delayed by the time to develop severe diseases; and 3) the number of new deaths due to Covid-19, biased by differential risk of death and the exact definition of a Covid-19 death, and further delayed by the time to death.

Each of these indicators provides a different view on the epidemic and therefore contains potentially useful information. However, any interpretation of their behaviour needs to reflect these biases and lags and is best done in combination with the other indicators. One approach that allows this in a principled manner is to use the different data sets to separately track the time-varying reproduction number, *R*_*t*_, the average number of secondary infections generated by each new infected person (5). Because *R*_*t*_ quantifies changes in infection levels, it is independent of the level of overall ascertainment as long as this does not change over time or is explicitly accounted for (6). At the same time, the underlying observations in each data source may result from different lags from infection to observation. However, if these delays are correctly specified then transmission behaviour over time can be consistently compared via estimates of *R*_*t*_.

Different methods exist to estimate the time-varying reproduction number, and in the UK a number of mathematical and statistical methods have been used to produce estimates used to inform policy (7– 9). Empirical estimates of *R*_*t*_ can be achieved by estimating time-varying patterns in transmission events from mapping to a directly observed time-series indicator of infection such as reported symptomatic cases. This can be based on the probabilistic assignment of transmission pairs (10), the exponential growth rate (11), or the renewal equation (12,13). Alternatively, *R*_*t*_ can be estimated via mechanistic models which explicitly compartmentalise the disease transmission cycle into stages from susceptible through exposed, infectious, and recovered (14,15). This can include accounting for varying population structures and context-specific biases in observation processes, before fitting to a source of observed cases. Across all methods, key parameters include the time after an infection to the onset of symptoms in the infecting and infected, and the source of data used as a reference point for earlier transmission events (16,17).

In this study, we used a modelling framework based on the renewal equation, adjusting for delays in observation to estimate regional and national reproduction numbers of SARS-Cov-2 across England. The same method was repeated for each of three sources of data that are available in real time. After assessing differences in *R*_*t*_ estimates by data source, we explored why this variation may exist. We compared the divergence between *R*_*t*_ estimates with spatio-temporal variation in case detection, and the proportion at risk of severe disease, represented by the age distribution of test positive cases and hospital admissions and the proportion of deaths in care homes.

## Methods

### Data management

Three sources of data provided the basis for our *R*_*t*_ estimates. Time-series case data were available by specimen date of test. This was a de-duplicated dataset of Covid-19 positive tests notified from all National Health Service (NHS) settings (Pillar One of the UK Government’s testing strategy)(18) and by commercial partners in community settings outside of healthcare (Pillar Two). Hospital admissions were also available by date of admission if a patient had tested positive prior to admission, or by the day preceding diagnosis if they were tested after admission. Death data were available by date of death and included only those which occurred within 28 days of a positive Covid-19 test in any setting. All data were publicly available and taken from the UK government source (19,20), and were aggregated to the seven English regions used by the NHS.

To provide context for *R*_*t*_ estimates, we sourced weekly data on regional and national test positivity (percentage positive tests of all tests conducted) from Public Health England (21), available as weekly average percentages from 10th May. From the same source, we also identified the age distributions of cases admitted to hospital and all test-positive cases. Hospital admissions by age were available as age bands with rates per 100,000, so we used regional population data from 2019 (22) to approximate the raw count. We separately sourced daily data on the number of deaths in care homes by region from March, available from 12th April (23). Care homes are defined as supported living facilities (residential homes, nursing homes, rehabilitation units and assisted living units). Data were available by date of notification, which included an average 2-3 day lag after the date of death. We also drew on a database which tracked Covid-19 UK policy updates by date and area (24).

### R_t_ estimation

We estimated *R*_*t*_ using EpiNow2 version 1.2.0, an open-source package in R (13,25,26). This package implements a Bayesian latent variable approach using the probabilistic programming language Stan (27). To initialise the model, infections were imputed prior to the first observed case using a log linear model with priors based on the first week of observed cases. This means that the initial observations both inform the initial parameters and are then also fit, which makes the initial Rt estimates less reliable than later estimates. This was a pragmatic choice to allow the model to be identifiable when only estimating part of the observed epidemic. We explored other parameterisations, but these suffered from poor model identification. For each subsequent time step with observed cases, new infections were imputed using the sum of previous modelled infections weighted by the generation time probability mass function, and combined with an estimate of *R*_*t*_, to give the prevalence at time *t* (12). The generation time was assumed to follow a gamma distribution that was fixed over time but varied between samples, with priors drawn from the literature for the mean and standard deviation (28).

These infection trajectories were mapped to reported case counts (*D*_*t*_) by convolving over an incubation period distribution and report delay distribution (ξ). We assumed a negative binomial observation model for observed reported case counts (*C*_*t*_), with overdispersion □ using an exponential prior with mean 1 and mean *D*_*t*_. We combined this with a multiplicative day of the week effect (ω(tmod7)) with an independent effect for each day of the week. We controlled temporal variation using an approximate Gaussian process (29) with a squared exponential kernel (GP).

In mathematical notation:

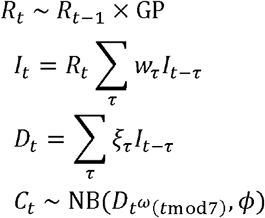

The length scale and magnitude of the kernel were estimated during model fitting. We used an inverse gamma prior for the length scale, optimising shape and scale values to give a distribution with 98% of the density between 2 and 21 days, and the prior on the magnitude was standard normal. Each region was fitted independently using Markov-chain Monte Carlo (MCMC). Eight chains were used with a warmup of 1,000 samples and 2,000 samples post warmup. Convergence was assessed using the R hat diagnostic.

We used a gamma distributed generation time with mean 3.6 days (standard deviation (SD) 0.7), and SD of 3.1 days (SD 0.8), sourced from (28). Instead of the incubation period used in the original study (which was based on fewer data points), we refitted using a log-normal incubation period with a mean of 5.2 days (SD 1.1) and SD of 1.52 days (SD 1.1)(30). This incubation period was also used to convolve from unobserved infections to unobserved symptom onsets (or a corresponding viral load in asymptomatic cases) in the model. When fitting the model, the time interval distributions had independent priors placed on the mean and standard deviation of their respective log-normal distributions.

We estimated both the delay from onset to positive test (either in the community or in hospital) and the delay from onset to death as log-normal distributions using a subsampled Bayesian bootstrapping approach (with 100 subsamples each using 250 samples) from given data on these delays. Our delay from date of onset to date of positive test (either in the community or in hospital) was taken from a publicly available linelist of international cases (31). We removed countries with outlying delays (Mexico and the Philippines). The resulting delay data had a mean of 4.4 days and standard deviation (SD) 5.6. Delays for hospital admissions and test positives were treated as having the same delay from infection to onset and observation. For the delay from onset to death we used data taken from a large observational UK study (32). We re-extracted the delay from confidential raw data, with a mean delay of 14.3 days (SD 9.5). There was insufficient data available on the various reporting delays to estimate spatially- or temporally-varying delays, so they were considered to be static over the course of the epidemic, although we discuss the effects of this assumption. We have also discussed this approach more extensively in (25).

### Comparison of R_t_ estimates

We compared *R*_*t*_ estimates by data source, plotting each by region over time. To avoid the first epidemic wave obscuring visual differences, all plots were limited to the earliest date that any *R*_*t*_ estimate for England crossed below 1 after the peak. We also identified the time at which each *R*_*t*_ estimate fell below 1, the local minima and maxima of median *R*_*t*_ estimates, and the number of times in the time-series that each *R*_*t*_ estimate crossed its own median, before comparing these across regions and against the total count of the raw data.

We investigated correlations between *R*_*t*_ estimates and the demographic and social context of transmission. We used linear regression to assess whether the level of raw data count influenced oscillations in *R*_*t*_. We assessed the influence of local outbreaks using test positivity. We used a 5% threshold for positivity as the level at which testing is either insufficient to keep pace with widespread community transmission (33), or where outbreaks have already been detected and tests targeted to those more likely to be positive. We plotted this against raw data and *R*_*t*_, and also used linear regression to test the association. We interpreted results in light of known outbreaks and policy changes. We plotted and qualitatively assessed variation in *R*_*t*_ estimates against the age distribution of cases over time, and similarly explored patterns in *R*_*t*_ estimates against the qualitative proportion of cases to all deaths. The latter was not assessed quantitatively due to differences in reference dates (23). With the exception of fitting the delay from onset to death (held confidentially), code and data to reproduce this analysis is available (34).

## Results

Across England, the Covid-19 epidemic peaked at 4,798 reported test-positive cases (on the 22^nd^ April), 3,099 admissions (1^st^ April), and 975 deaths (8^th^ April) per day (Figure 1A). Following the peak, a declining trend continued for daily counts of admissions and deaths, while daily case counts from all reported test-positive cases increased from July and had more than tripled by August (from 571 on 30th June to 1,929 on 1^st^ September). Regions followed similar patterns over time to national trends. However, in the North East and Yorkshire, Midlands, and North West, incidence of test-positive cases did not decline to near the count of admissions as in other regions, and also saw a small temporary increase during the overall rise in case counts in early August.

**Figure 1.**
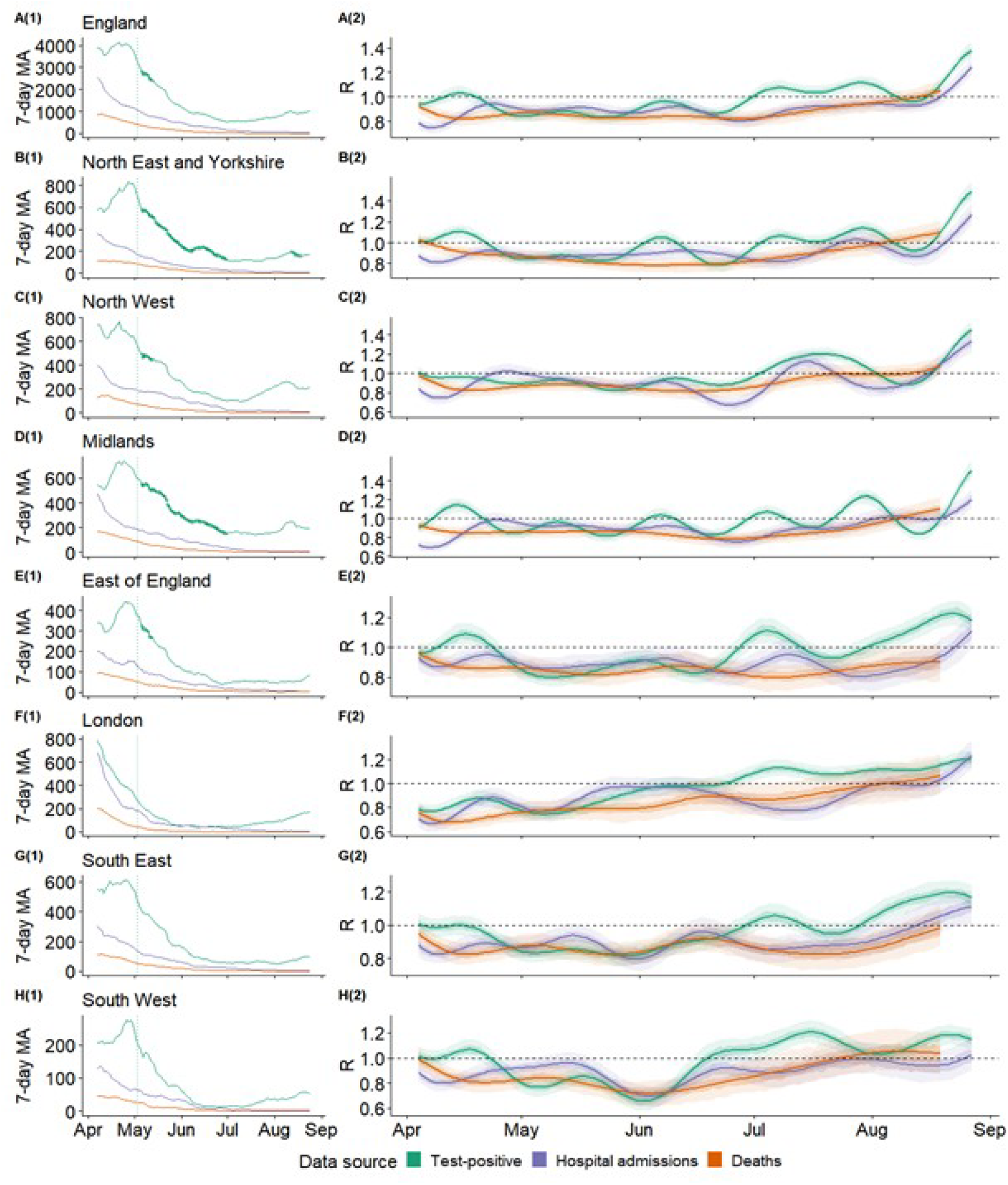
Epidemic dynamics across **(A)** England and **(B-H)** seven English National Health Service regions, 5th April through 27th August 2020. **A1-H1**: Daily counts of confirmed cases by data source, as centred seven day moving average. Counts marked with crosses indicate dates within weeks which averaged >5% test positivity (positive / all tests per week). Vertical dotted line indicates the start of national mass community testing on 3rd May. **A2-H2**: Estimates of Rt, (median, with 50% (darker shade) and 90% (lightest shade) credible interval), derived from each data source. Data sources include all test-positive cases, hospital admissions, and deaths with a positive test in the previous 28 days.

Following the initial epidemic peak in mid-March, the date at which *R*_*t*_ estimates crossed below 1 varied by both data source and geography (Figure 1B, Figure 2). The first region to cross into a declining epidemic was London, on the 26^th^ March according to an *R*_*t*_ estimated from deaths (where the lower 90% CrI crossed below 1 on the 24th and the upper CrI on the 28^th^ March). However, the data source used to estimate *R*_*t*_ was as important as any regional variation in estimating the earliest date of epidemic decline. *R*_*t*_ estimated from hospital admissions gave the earliest estimate of a declining epidemic, while using all test-positive cases to estimate *R*_*t*_ took the longest time to reach a declining epidemic, in all but one region (East of England). This difference by data source varied by up to 21 days in the North East and Yorkshire, where hospital admissions gave a median *R*_*t*_ estimate under 1 on the 1^st^ April (90%CrIs 31^st^ March, 2^nd^ April), but the median *R*_*t*_ estimate from test-positive cases crossed 1 on only the 22^nd^ April (90%CrIs 1^st^ April, 25^th^ April).

**Figure 2.**
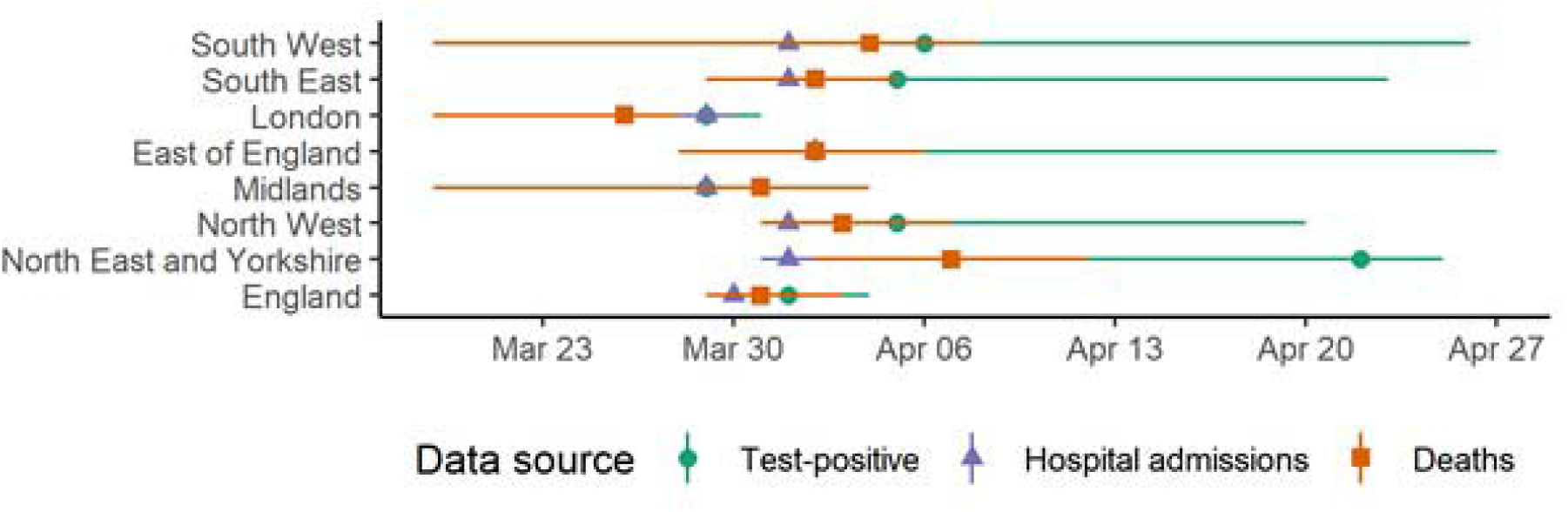
Dates on which Rt estimate crossed 1 after first epidemic peak, median and 90% credible interval, by data source for England and seven NHS regions.

When not undergoing a clear state change, *R*_*t*_ estimates from all data sources oscillates, with oscillations damped when *R*_*t*_ estimates were transitioning to new levels. In England and all NHS regions, test-positive cases showed evidence of larger damped oscillations from July when a state change occurred to *R*_*t*_ over 1. In England, *R*_*t*_ estimates from test-positive cases increased from 0.99 (90%CrI 0.94-1.04) on 30^th^ June to 1.37 (90%CrI 1.31-1.1.44) on 27^th^ August. Meanwhile, the timing and duration of oscillations did not align between *R*_*t*_ estimates (Figure 1B). In some regions, the difference between *R*_*t*_ estimates was consistent over time, such as between *R*_*t*_ from admissions and deaths in the South East. In other regions such as the Midlands this was not the case, with the divergence between the *R*_*t*_ estimates from test-positive cases, admissions, and deaths each varying over time. *R*_*t*_ estimates from test-positive cases were the most likely to differ from estimates derived from other data sources across all regions. Across all regions, *R*_*t*_ estimates from deaths had slower damped oscillations compared to estimates from test-positive cases or hospital admissions. However, oscillations in *R*_*t*_ estimates did not appear to be linked to the level of raw data counts in each source (SI Figure 2).

More rapid oscillations in *R*_*t*_ estimates from test-positive cases appeared to be linked to targeted testing of case clusters, seen in high test positivity (Table SI2). Both the North East and Yorkshire and the Midlands saw more frequent oscillations in *R*_*t*_ estimates from test-positive cases than other regions. The *R*_*t*_ estimates from cases crossed its own median 10 times over the time-series in both regions, while in all other NHS regions this averaged 6 times, and oscillations in *R*_*t*_ estimates from cases also had a shorter duration in the North East and Yorkshire and the Midlands compared to other regions (Table SI1). Across all regions, 84% of weeks with over 5% positivity (N=19) were in the North East and Yorkshire and the Midlands (Figure 2A). In these regions, positivity peaked on the week of 9^th^ May at 14% and 12% respectively, and overall averaged 6% (95%CI 4.4-7.6%) and 5.9% (95%CI 4.6-7.2%, weeks of 10^th^ May to 22^nd^ August) respectively. High test positivity is likely to have resulted from targeted testing among known local outbreaks in these regions. In the Midlands, these included local restrictions and increased testing across Leicester and in a Luton factory (restrictions between 4^th^ and 25^th^ July (35)). In Yorkshire case clusters were detected with local restrictions in Bradford, Calderdale, and Kirklees (with restrictions from 5 August (36)).

In England, a divergence between *R*_*t*_ estimated from cases versus *R*_*t*_ estimated from deaths and admissions coincided with a decline in the age distribution among all test-positive cases in England to a younger population (Figure SI2A). From mid-April to June, national estimates of *R*_*t*_ from test-positive cases remained around the same level as those from admissions or deaths, while after this, cases diverged to a higher steady state (Figure 1A). On the 23^rd^ May, the median *R*_*t*_ estimated from cases matched that of deaths at 0.83 (both with 90% CrIs 0.78-0.89), but this was followed by a 78 day period before the two estimates were again comparable, on 8^th^ August. Over this period the median *R*_*t*_ estimate from cases was on average 14% higher (95%CI 12-15%). Meanwhile, the share of test-positive cases under age 50 increased from under one-quarter of cases in the week of 28^th^ March (24%, N=16,185), to accounting for nearly three-quarters of cases by 22^nd^ August (77%, N=6,733). While the percentage of test-positive cases aged 20-49 increased consistently from April to August, the 0-19 age group experienced a rapid increase over mid-May through July, increasing by a mean 1% each week over May 9^th^ through August 1^st^ (from 4% of 18,774 cases to 14.8% of 5,017 cases).

Similarly, *R*_*t*_ estimates from admissions in England oscillated over June through July, potentially linked to the age distribution of hospital admissions. From 0.92 (90%CrI 0.87-0.98) on the 11^th^ June, *R*_*t*_ estimated from admissions fell to 0.8 (90%CI 0.75-0.85)) on the 27^th^ June. In contrast, this transition was not observed in the *R*_*t*_ estimate based on test-positive cases (Figure 1A). Older age groups dominated Covid-19 hospital admissions, where 0-44 years never accounted for more than 12.8% of hospital-based cases (a maximum in the week of 22^nd^ August, N=690; Figure SI2B). While the proportion of hospital admissions aged 75+ remained steady over May through mid-June, this proportion appeared to oscillate over July through August (standard deviation of weekly percentage at 6.1 over June-August, compared to 5.4 in months March-May). These variations were not seen in the proportion aged 70+ in the test-positive case data, which saw a continuous decline from 30% at the start of June to 7% by August.

*R*_*t*_ estimated from either admissions or deaths experienced near-synchronous local peaks across regions over April and May. We compared this *R*_*t*_ estimated from deaths with its source data and a separate regional dataset of deaths in care homes. In the South East and South West, the *R*_*t*_ estimates from deaths rose over April, with a peak in early May. In the South West, the median *R*_*t*_ estimate from deaths increased by 0.04 from 22^nd^ April to 7^th^ May (from 0.8 (90%CrI 0.72-0.88) to 0.84 (90%CrI 0.76-0.95)); and by 0.06 from the 17^th^ April to 4^th^ May in the South East (from 0.82 (90%CrI 0.77-0.9) to 0.88 (90%CrI 0.72-0.88)). In both these regions, this early May peak in *R*_*t*_ estimates from deaths coincided with similarly rising *R*_*t*_ estimates from hospital admissions, while the reverse trend was seen in *R*_*t*_ estimates from cases. In all regions, care home deaths peaked over the 22^nd^-29^th^ April (by date of notification; Figure SI3). This was later than regional peaks in the raw count of all deaths in any setting (which peaked between the 8^th^-16^th^ April, by date of death), even accounting for a 2-3 day reporting lag. This meant that the proportion of deaths from care homes varied over time, where in the South East and South West, deaths in care homes appeared to account for nearly all deaths for at least the period mid-May to July.

### Discussion

We estimated the time-varying reproduction number for Covid-19 over March through August across England and English NHS regions, using test-positive cases, hospital admissions, and deaths with confirmed Covid-19. Our estimates of transmission potential varied for each of these sources of infections, and the divergence between estimates from each data source was not consistent within or across regions over time, although estimates based on hospital admissions and deaths were more spatio-temporally synchronous than compared to estimates from cases. We compared differences in *R*_*t*_ estimates to the extent and context of transmission and found that the difference between *R*_*t*_ estimated from cases, admissions, and deaths may be linked to uneven rates of testing, the changing age distribution of cases, and outbreaks in care home populations.

*R*_*t*_ estimates varied by data source, and the extent of variation itself differed by region and over time. Following the initial epidemic peak in mid-March, the date at which *R*_*t*_ estimates crossed below 1 varied by both data source and geography, following which *R*_*t*_ estimates from all data sources varied when not undergoing a clear state change. The differences in these oscillations by data source may indicate different underlying causes. This implies that each data source was influenced differently by changes in subpopulations over time.

Increasingly rapid oscillations in *R*_*t*_ estimates from test-positive cases were associated with higher test-positivity rates. Increasing test-positivity rates could be an indication of inconsistent community testing, with the observation of an initial rise in transmission amplified by expanded testing and local interventions where a cluster of new, mild cases has been identified (18). This targeted testing may drive regionally localised instability in case detection and resulting *R*_*t*_ estimates but may not reflect changes in underlying transmission. This is a limitation of monitoring epidemic dynamics using test-positive surveillance data in areas where testing rates vary across the population and over time. This also suggests that *R*_*t*_ estimates from admissions may be more reliable than that from all test-positive cases for indicating the relative intensity of an epidemic over time (37).

We hypothesised that variations in *R*_*t*_ estimates were also related to changes in the age distribution of cases over time, because age is associated with severity (38,39). If each data source represented a different sample of this age-severity gradient, and transmission also varied by age or severity, *R*_*t*_ estimates from each source would diverge. Early in the epidemic, tests were largely limited to hospital settings, and disproportionately represented healthcare workers compared to the general population. This sampling bias would be reflected in the *R*_*t*_ from test-positive cases. The early peak in *R*_*t*_ could then represent a substantial separate route of transmission in healthcare settings, in a wave of nosocomial infections (40). If healthcare workers were less susceptible to severe disease than those older than working age, an early peak in *R*_*t*_ estimated from test-positive cases would not have been represented in *R*_*t*_ estimated from hospital admissions or deaths. Meanwhile, either hospital admissions or deaths data would be more representative of sampling a separate route of transmission among the general population. If infections spread through the general population later than nosocomial infections, then the timing of peaks in *R*_*t*_ estimates from each data source would not have matched.

From late spring, outbreaks in care homes may have contributed to a divergence between Rt estimates from test positive cases and other data sources. All regions saw a near-synchronous local peak in *R*_*t*_ estimated from hospital admissions over spring, which was not seen in *R*_*t*_ estimated from test-positive cases. This may have reflected the known widespread regional outbreaks in care homes. The care home population is on average older and more clinically vulnerable than the general population, while also being less likely to appear for community testing (41,42). Increased transmission in care homes would then be seen in an increased *R*_*t*_ from hospital admissions, but not observed in an *R*_*t*_ from test-positive cases.

Similarly, the age-severity gradient may have impacted transmission estimates later in the epidemic when community testing became more widely available. We found that from June onwards, *R*_*t*_ estimates from all test-positive cases appeared to increasingly diverge away from *R*_*t*_ estimates from admissions and deaths, transitioning into a separate, higher, steady state. This was followed by the observed age distribution of all test-positive cases becoming increasingly younger, while the age distribution of admissions remained approximately level. Because of the severity gradient, this suggested the *R*_*t*_ estimates from all test-positive cases and admissions were more biased by the relative proportion of younger cases and older cases respectively than the *R*_*t*_ estimates from admissions or deaths.

Our analysis was limited where data or modelling assumptions did not reflect underlying differences in transmission. *R*_*t*_ estimates can become increasingly uncertain and unstable with lower case counts. Further, estimated unobserved infections were mapped to reported cases or deaths using two delay distributions: the time from infection to test in the community or hospital, and a longer delay from infection to death. Mis-specification of the priors would have created bias in the temporal distribution of all resulting *R*_*t*_ estimates, with estimated dates of infection and *R*_*t*_ incorrectly shifted too much or little in time compared to the true infection curve, and decreased accuracy of *R*_*t*_ estimates (43).

We used the same distribution priors for both delays after symptom onset to positive test, and to hospital admission. This may be inaccurate where cases with mild symptoms take longer to present for testing than severe cases presenting for hospital admission, or vice versa. The difference between the two delays over time may also have varied, with a possible decrease in delay to reported tests when mass community testing became available over the summer. This would have had a differential impact on the accuracy of *R*_*t*_ estimates over time in either direction, which could explain some of the oscillations in *R*_*t*_ estimates from test-positive case data compared to hospital admissions. We had no data over time on delays from symptom onset to reporting in each data source with which to test this hypothesis. However, we have mitigated some of the impact of this by using a sub-sampled bootstrap of the available delay data when estimating the delay distribution priors. This inflated the uncertainty of these priors in line with the hypothesis that they varied over time. This adjustment may be conservative if the delay distributions are stable over time.

Spatial dependence in delay distributions may also have contributed to their mis-specification and increased uncertainty in *R*_*t*_ estimates. We observed that the variation in *R*_*t*_ estimates from admissions and deaths often showed comparable levels and patterns in oscillations over time but were out of phase with each other. This may have been due to using data sources from different populations for each delay estimate. To estimate the delay between symptom onset to either a positive test or hospitalisation, we used a linelist of all patients publicly reported globally, which had a mean delay 5.4 days (SD 5.6). This varied only slightly from an early estimate in the UK epidemic, where the delay from onset to hospitalisation had a mean 5.14 days (SD 4.2) in confidential Public Health England (FF100) data (44). Meanwhile, the same global public linelist contained few records with delay from onset to death, with mean 11.4 (SD 16.5). We compared this to confidential UK data from an observational study which had mean delay 14.3 days (SD 9.5) (32).

Comparing each type and source of delay, we judged the benefits of using open data to outweigh the minor observed spatial variation of the delay from onset to test or admission, although at the expense of increased uncertainty. However, we judged the difference in delay from onset to death in the UK compared to public (international) data was sufficiently meaningful to justify using confidential UK data in order to maintain accuracy of the *R*_*t*_ estimate from deaths. The difference in geographic source of delay distributions should not have substantially altered our conclusions about discrepancies between central estimates of *R*_*t*_ from either test-positives or admissions, compared to *R*_*t*_ estimated from deaths. However, using the international public linelist for the delay to test or admission may have introduced additional uncertainty around the respective *R*_*t*_ estimates, compared to greater accuracy (reduced uncertainty) in estimates of *R*_*t*_ from deaths based on a UK-specific delay distribution.

The data sources themselves may also have been inaccurate or biased, which would change the representation of the population we have assumed here. For example, we excluded data from other nations of the UK (Wales, Scotland and Northern Ireland) in our analysis, as these differed in both availability over time and in data collection and reporting practices (19,45). English regional data may also contain bias where new parts of the population might be under focus for testing efforts, or the population characteristics of hospital admissions from Covid-19 may have changed over time with changes in clinical criteria or hospital capacity for admission. This would mean that an *R*_*t*_ estimate from these data sources would represent different source populations over time, limiting our ability to reliably compare against *R*_*t*_ estimates from other data sources. Where possible we highlighted this by comparing *R*_*t*_ estimates to known biases and changes in case detection and reporting.

Our approach is unable to make strong causal conclusions about varying transmission, and assumptions about sampling and the representation of subpopulations remain implicit. Alternatively, varying epidemics in subpopulations could have been addressed with mechanistic models that explicitly consider transmission in different settings and are fitted to multiple data sources. However, these require additional assumptions, detailed data to parameterise, and may be time-consuming to develop. In the absence of data, the number of assumptions required for these models can introduce inherent structural biases. Our approach contains few structural assumptions and therefore may be more robust when data are sparse, or information is required in real-time.

We conclude that when estimating *R*_*t*_, the choice of data source should be guided by the policy context in which the estimates will be used and interpreted. This work highlights that there is no clear superior choice of data source, while *R*_*t*_ estimates are sensitive to assumptions about the underlying population of each data source. This means that both producers and users of *R*_*t*_ estimates should understand relevant biases in the data source’s population sampling strategy, such as by community case detection or patient severity, before drawing conclusions about transmission in the population as a whole.

We also recommend presenting concurrent *R*_*t*_ estimates jointly, rather than pooling estimates of *R*_*t*_ from different data sources. Pooling estimates would both suffer from unclear weighting and lose useful information about variation in subpopulation transmission. Although the reconstruction of the underlying transmission process from the reporting processes is robust, it is unclear how weights would be assigned based on likelihood to estimates from different data sources. Further, the variation in concurrent *R*_*t*_ estimates provides more information about population transmission than any single estimate, when considered in light of the sampling biases of each data source. This additional information can be useful to identify transmission intensity by subpopulation where access to high quality disaggregated data may not be available in real time. While this can be difficult to interpret without specific knowledge of population structure and dynamics, this information would be lost altogether in a single or pooled estimate of *R*_*t*_. In contrast, if policy were to be based on either a single or an averaged *R*_*t*_ estimate, it would be unclear what any recommendation should be and for whom.

Future work could explore systematic differences in the influence of data sources on *R*_*t*_ estimate by extending the comparison of *R*_*t*_ by data source to other countries or infectious diseases. Additionally, work should also clarify the potential for comparing *R*_*t*_ estimates in real-time tracking of outbreaks and explore the inconsistencies in case detection over time and space, where a cluster of cases leads to a highly localised expansion of community testing, creating an uneven spatial bias in transmission estimates. These findings may be used to improve *R*_*t*_ estimation and identify findings of use for epidemic control. Based on the work presented here we now provide *R*_*t*_ estimates, updated each day, for test positive cases, admissions, and deaths in each NHS region and in England. Our estimates are visualised on our website, are available for download, and are produced using publicly accessible code (46,47).

Tracking differences by data source can improve understanding of variation in testing bias in data collection, highlight outbreaks in new subpopulations and indicate differential rates of transmission among vulnerable populations, and clarify the strengths and limitations of each data source. Our approach can quickly identify such patterns in developing epidemics that might require further investigation and early policy intervention. Our method is simple to deploy and scale over time and space using existing open-source tools, and all code and estimates used in this work are available to be used or re-purposed by others.

## Data Availability

The code that supports the findings of this study is available on Github, DOI: 10.5281/zenodo.4029075. Other sources of data were derived from the linked resources available in the public domain.

https://github.com/epiforecasts/rt-comparison-uk-public

https://github.com/beoutbreakprepared/nCoV2019/tree/master/latest_data

https://www.health.org.uk/news-and-comment/charts-and-infographics/covid-19-policy-tracker

https://www.ons.gov.uk/peoplepopulationandcommunity/populationandmigration/populationestimates/datasets/populationestimatesforukenglandandwalesscotlandandnorthernireland

https://www.gov.uk/government/publications/national-covid-19-surveillance-reports.

https://www.ons.gov.uk/peoplepopulationandcommunity/birthsdeathsandmarriages/deaths/datasets/numberofdeathsincarehomesnotifiedtothecarequalitycommissionengland

## Acknowledgements

The following authors were part of the Centre for Mathematical Modelling of Infectious Disease COVID-19 Working Group. Each contributed in processing, cleaning and interpretation of data, interpreted findings, contributed to the manuscript, and approved the work for publication: Fiona Yueqian Sun, C Julian Villabona-Arenas, Emily S Nightingale, Alicia Showering, Gwenan M Knight, Yang Liu, Kaja Abbas, Akira Endo, Alicia Rosello, Rachel Lowe, Matthew Quaife, Amy Gimma, Oliver Brady, Nicholas G. Davies, Anna Vassal, W John Edmunds, Jack Williams, Simon R Procter, Rosalind M Eggo, Yung-Wai Desmond Chan, Rosanna C Barnard, Georgia R Gore-Langton, Naomi R Waterlow, Charlie Diamond, Timothy W Russell, Graham Medley, Katherine E. Atkins, Kiesha Prem, David Simons, Megan Auzenbergs, Damien C Tully, Christopher I Jarvis, Kevin van Zandvoort,Carl A B Pearson, Thibaut Jombart, Anna M Foss, Adam J Kucharski, Billy J Quilty, Hamish P Gibbs, Samuel Clifford, Petra Klepac.

## Funding

The following funding sources are acknowledged as providing funding for the named authors. Wellcome Trust (210758/Z/18/Z: JDM, JH, KS, NIB, SA, SFunk, SRM). This research was partly funded by the Bill & Melinda Gates Foundation (INV-003174: MJ). This project has received funding from the European Union’s Horizon 2020 research and innovation programme - project EpiPose (101003688: MJ). The following funding sources are acknowledged as providing funding for the working group authors. Alan Turing Institute (AE). BBSRC LIDP (BB/M009513/1: DS). This research was partly funded by the Bill & Melinda Gates Foundation (INV-001754: MQ; INV-003174: KP, MJ, YL; NTD Modelling Consortium OPP1184344: CABP, GFM; OPP1180644: SRP; OPP1183986: ESN; OPP1191821: KO’R, MA). BMGF (OPP1157270: KA). Foreign, Commonwealth and Development Office (FCDO)/Wellcome Trust (Epidemic Preparedness Coronavirus research programme 221303/Z/20/Z: CABP, KvZ). DTRA (HDTRA1-18-1-0051: JWR). Elrha R2HC/UK FCDO/Wellcome Trust/This research was partly funded by the National Institute for Health Research (NIHR) using UK aid from the UK Government to support global health research. The views expressed in this publication are those of the author(s) and not necessarily those of the NIHR or the UK Department of Health and Social Care (KvZ). ERC Starting Grant (#757699: JCE, MQ, RMGJH). This project has received funding from the European Union’s Horizon 2020 research and innovation programme - project EpiPose (101003688: KP, MJ, PK, RCB, WJE, YL). This research was partly funded by the Global Challenges Research Fund (GCRF) project ’RECAP’ managed through RCUK and ESRC (ES/P010873/1: AG, CIJ, TJ). HDR UK (MR/S003975/1: RME). MRC (MR/N013638/1: NRW). Nakajima Foundation (AE). NIHR (16/136/46: BJQ; 16/137/109: BJQ, CD, FYS, MJ, YL; Health Protection Research Unit for Immunisation NIHR200929: NGD; Health Protection Research Unit for Modelling Methodology HPRU-2012-10096: TJ; NIHR200929: FGS, MJ; PR-OD-1017-20002: AR, WJE). Royal Society (Dorothy Hodgkin Fellowship: RL; RP\EA\180004: PK). UK DHSC/UK Aid/NIHR (ITCRZ 03010: HPG). UK MRC (LID DTP MR/N013638/1: GRGL, QJL; MC_PC_19065: AG, NGD, RME, SC, TJ, WJE, YL; MR/P014658/1: GMK). Authors of this research receive funding from UK Public Health Rapid Support Team funded by the United Kingdom Department of Health and Social Care (TJ). Wellcome Trust (206250/Z/17/Z: AJK, TWR; 206471/Z/17/Z: OJB; 208812/Z/17/Z: SC; 208812/Z/17/Z: SFlasche). No funding (AKD, AMF, AS, CJVA, DCT, JW, KEA, SH, YJ, YWDC)

## Notes

### Competing Interest Statement

The authors have declared no competing interest.

### Author Declarations

Ethical approval was not required for this study, which used anonymised public secondary data sources only.

### Summary of Updates

Corrected typesetting and clarified the paper title. Added detail in methods including full specification of model, handling of uncertainty with clarification of priors and delay and generation time distribution. Additional discussion of spatial and temporal dependence in delay distributions, nosocomial outbreaks, and recommendation against pooling estimates.

