## Supplementary Figures 1-3, Supplemental Tables 1-2 for "Exploring surveillance data biases when estimating the reproduction number: with insights into subpopulation transmission of Covid-19 in England"

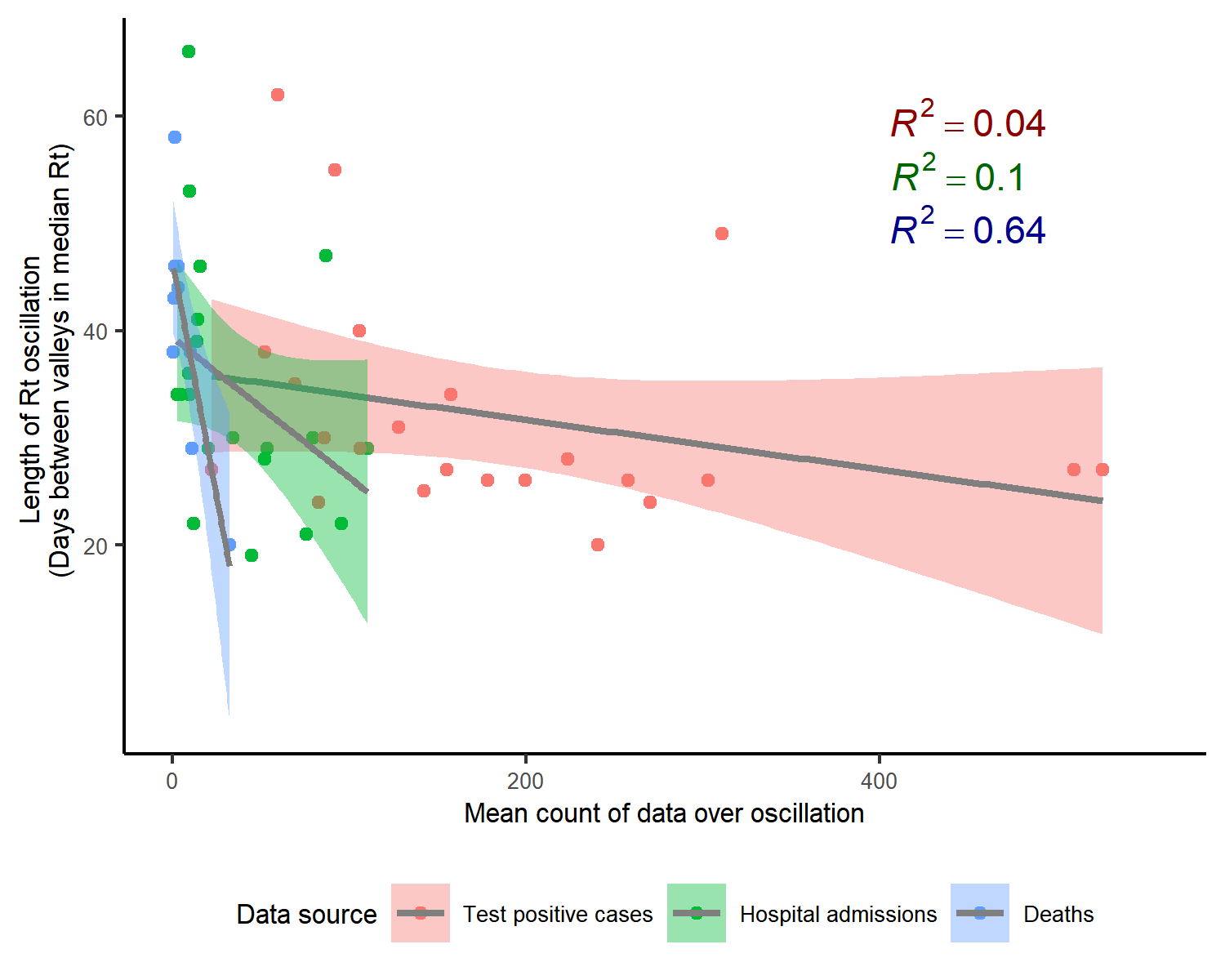

*Figure SI1. Duration of each R_t_ oscillation against the mean of data over the oscillation. Shown with fitted linear models by data source. Scatter points represent each oscillation across 7 English NHS regions.* *For R_t_ derived from deaths, longer durations of oscillations appeared to be related to lower raw counts (coefficient: -0.88, R2 0.64), but this had a very small sample size (N oscillations = 9) and this relationship was not meaningful for cases or hospital admissions.*

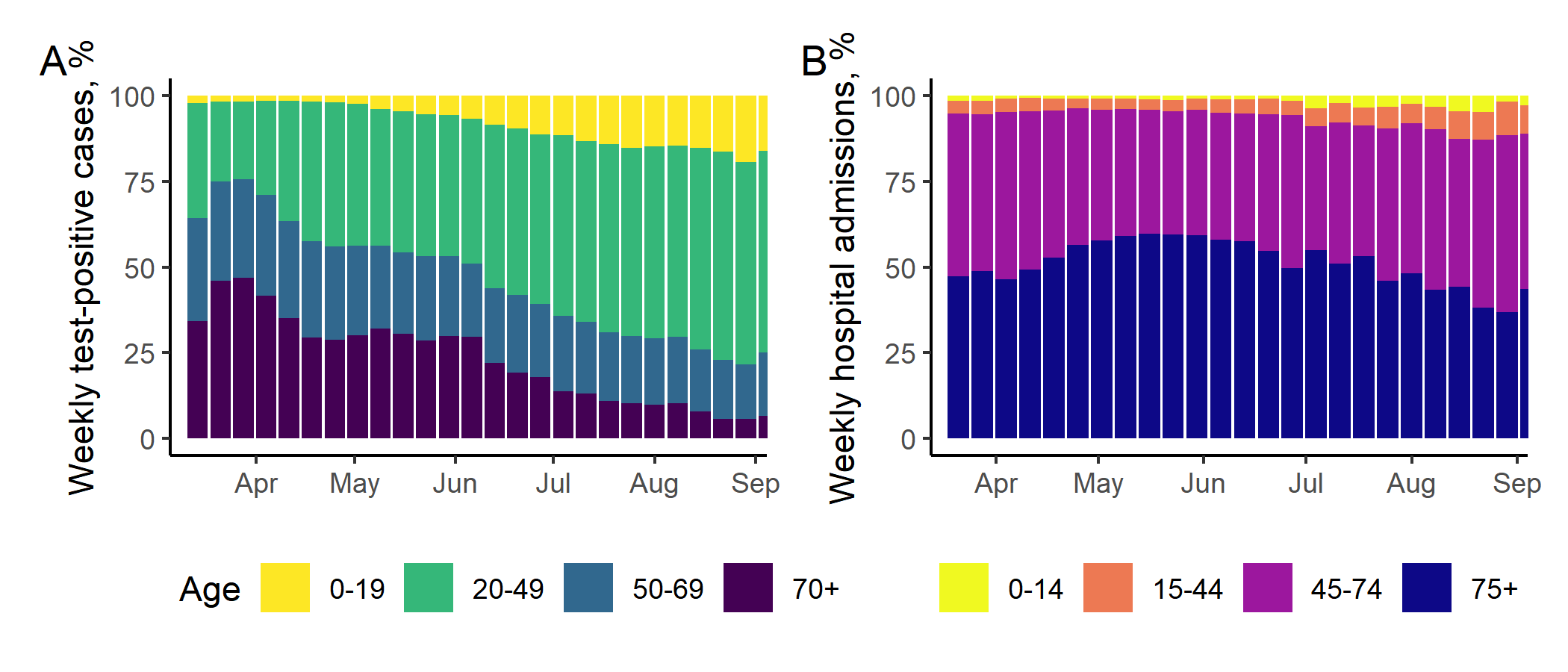

*Figure SI2. Weekly percentage of cases in England by age, among all test-positive cases (A) and newly diagnosed hospital admissions (B).*

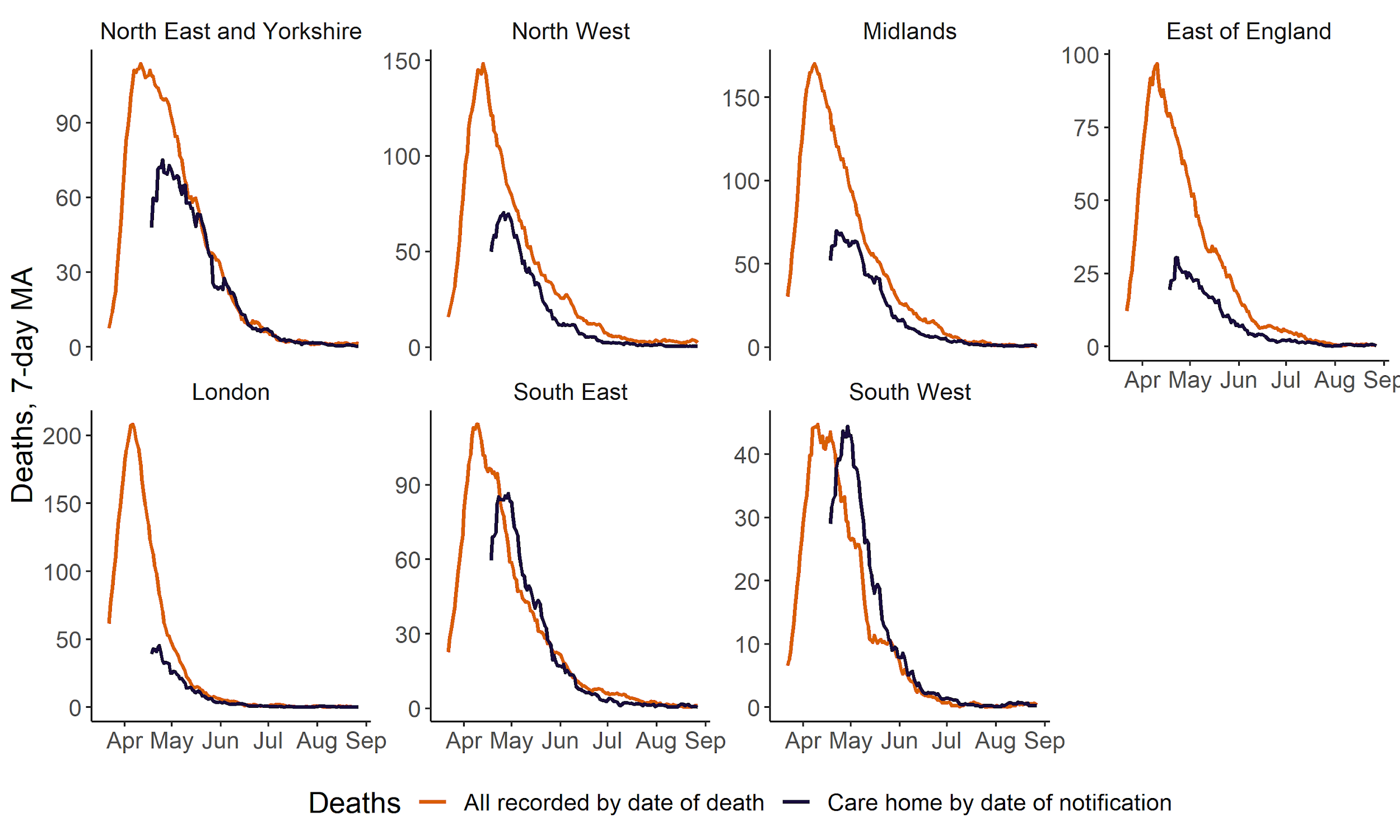

*Figure SI3. Deaths in all settings by date of death, and deaths in care homes by date of notification to Clinical Quality Commission, shown by English NHS region, April to August 2020. Date of notification is typically 2-3 days after the date of death* [22]*. Counts are shown as a seven-day moving average. Care homes data reported from April 2020.*

| **Region** | **Data source for Rt estimation** | **Earliest date median Rt <1** | **Median Rt after first wave* (90% CrI)** | **Minimum median Rt after first wave* (date, median [90%CrI])** | **Maximum median Rt after first wave* (date, median [90%CrI])** | **Number of days where median Rt crossed series median** | **Mean number of days between troughs** (mean, 95% CI)** |
| --- | --- | --- | --- | --- | --- | --- | --- |
| **England** | *Test-positive cases* | 01 Apr | 0.97 (0.85-1.11) | 23 May (0.83 [0.78-0.89]) | 27 Aug (1.37 [1.31-1.44]) | 6 | 32 (21-44, n=4) |
|  | *Hospital admissions* | 30 Mar | 0.9 (0.81-0.98) | 08 Apr (0.75 [0.7-0.79]) | 27 Aug (1.24 [1.17-1.31]) | 8 | 27 (24-29, n=3) |
|  | *Deaths* | 31 Mar | 0.86 (0.8-0.98) | 27 Jun (0.82 [0.76-0.89]) | 19 Aug (1.06 [0.97-1.14]) | 4 | 40 (n=1) |
| **North East and Yorkshire** | *Test-positive cases* | 22 Apr | 0.96 (0.82-1.13) | 22 Jun (0.79 [0.73-0.85]) | 27 Aug (1.49 [1.4-1.58]) | 10 | 26 (23-29, n=5) |
|  | *Hospital admissions* | 01 Apr | 0.89 (0.82-1.01) | 09 Apr (0.81 [0.76-0.87]) | 27 Aug (1.27 [1.16-1.38]) | 8 | 31 (23-39, n=4) |
|  | *Deaths* | 07 Apr | 0.86 (0.77-1.02) | 04 Jun (0.78 [0.72-0.85]) | 19 Aug (1.11 [1-1.21]) | 2 | NA |
| **North West** | *Test-positive cases* | 05 Apr | 0.94 (0.85-1.19) | 26 May (0.82 [0.76-0.87]) | 27 Aug (1.45 [1.37-1.54]) | 4 | 34 (22-46, n=3) |
|  | *Hospital admissions* | 01 Apr | 0.91 (0.74-1.09) | 25 Jun (0.67 [0.6-0.75]) | 27 Aug (1.33 [1.22-1.44]) | 8 | 38 (30-47, n=3) |
|  | *Deaths* | 03 Apr | 0.88 (0.8-1.02) | 15 Jun (0.81 [0.73-0.88]) | 19 Aug (1.08 [0.97-1.17]) | 4 | NA |
| **Midlands** | *Test-positive cases* | 29 Mar | 0.96 (0.84-1.17) | 23 May (0.82 [0.75-0.88]) | 27 Aug (1.51 [1.41-1.61]) | 12 | 27 (25-28, n=5) |
|  | *Hospital admissions* | 29 Mar | 0.91 (0.76-1.03) | 07 Apr (0.7 [0.64-0.75]) | 27 Aug (1.2 [1.11-1.31]) | 6 | 26 (22-30, n=4) |
|  | *Deaths* | 31 Mar | 0.86 (0.78-0.99) | 24 Jun (0.78 [0.71-0.85]) | 19 Aug (1.11 [1.01-1.23]) | 8 | 33 (12-54, n=2) |
| **East of England** | *Test-positive cases* | 02 Apr | 0.96 (0.82-1.15) | 09 May (0.8 [0.74-0.86]) | 22 Aug (1.23 [1.15-1.31]) | 6 | 36 (34-38, n=3) |
|  | *Hospital admissions* | 02 Apr | 0.89 (0.81-0.99) | 28 Jul (0.8 [0.71-0.9]) | 27 Aug (1.11 [0.99-1.24]) | 8 | 36 (28-45, n=3) |
|  | *Deaths* | 02 Apr | 0.85 (0.78-0.94) | 06 Jul (0.8 [0.71-0.88]) | 02 Apr (0.99 [0.93-1.05]) | 6 | 38 (24-51, n=2) |
| **London** | *Test-positive cases* | 29 Mar | 0.98 (0.77-1.15) | 07 May (0.74 [0.68-0.8]) | 25 Aug (1.2 [1.13-1.27]) | 4 | 35 (27-43, n=2) |
|  | *Hospital admissions* | 29 Mar | 0.89 (0.73-1.04) | 08 Apr (0.67 [0.6-0.73]) | 27 Aug (1.23 [1.12-1.36]) | 4 | 47 (16-78, n=2) |
|  | *Deaths* | 26 Mar | 0.84 (0.7-1.02) | 13 Apr (0.68 [0.61-0.75]) | 19 Aug (1.07 [0.91-1.24]) | 4 | 41 (36-46, n=2) |
| **South East** | *Test-positive cases* | 05 Apr | 0.96 (0.82-1.15) | 28 May (0.82 [0.75-0.87]) | 22 Aug (1.2 [1.14-1.27]) | 4 | 35 (19-51, n=3) |
|  | *Hospital admissions* | 01 Apr | 0.89 (0.81-1.02) | 31 May (0.8 [0.73-0.87]) | 27 Aug (1.11 [1-1.21]) | 8 | 29 (23-34, n=3) |
|  | *Deaths* | 02 Apr | 0.86 (0.79-0.95) | 25 May (0.82 [0.76-0.9]) | 19 Aug (0.99 [0.87-1.12]) | 6 | 48 (32-64, n=2) |
| **South West** | *Test-positive cases* | 06 Apr | 1.02 (0.73-1.18) | 02 Jun (0.66 [0.58-0.74]) | 16 Jul (1.21 [1.12-1.3]) | 6 | 39 (21-58, n=3) |
|  | *Hospital admissions* | 01 Apr | 0.9 (0.76-1.02) | 03 Jun (0.7 [0.61-0.78]) | 27 Aug (1.02 [0.91-1.18]) | 6 | 44 (28-59, n=2) |
|  | *Deaths* | 04 Apr | 0.85 (0.72-1.06) | 03 Jun (0.72 [0.62-0.81]) | 07 Aug (1.05 [0.9-1.24]) | 2 | 43 (n=1) |

*Table SI1. Key summary statistics for R_t_ from when R_t_ crossed 1, indicating the end of the first wave of the epidemic, to the most recent estimate. 90%CrI = credible interval (quantiles around median); 95%CI = confidence interval (standard error around mean)*

**End of first wave = earliest date Rt <1. Last estimate of Rt from cases and admissions: 28th August, from deaths: 19th August 2020*

***Troughs defined as local minima within a sequential fall and rise in the median Rt estimate*

| **Effect on regional average duration between troughs in Rt** | **Influence of % test positive** | | |
| --- | --- | --- | --- |
| Source of Rt | Intercept (95% CI) | Coefficient (95% CI) | Adjusted R^2^ |
| Test positive cases | 39 (36.7 to 42) | -2.2 (-2.8 to -1.4) | 0.87 (*p* <0.001) |
| Hospital admissions | 41 (29.5 to 52) | -2.1 (-5.5 to 1.2) | 0.18 (*p* 0.2) |
| Deaths | 45.4 (38 to 52.5) | -3.7 (-4.7 to 0.37) | 0.48 (*p* 0.07) |

*Table SI2**. Results of individual linear regressions of test positive on each Rt estimate from test-positive cases, hospital admissions, and deaths separately. Test positivity is the average % positive of all tests conducted. Data for seven English NHS regions and England (N=8).*
